# Tracking Smell Loss to Identify Healthcare Workers with SARS-CoV-2 Infection

**DOI:** 10.1101/2020.09.07.20188813

**Authors:** Julian J. Weiss, Tuki N. Attuquayefio, Elizabeth B. White, Fangyong Li, Rachel S. Herz, Theresa L. White, Melissa Campbell, Bertie Geng, Rupak Datta, Anne L. Wyllie, Nathan D. Grubaugh, Arnau Casanovas-Massana, M. Catherine Muenker, Ryan Handoko, Akiko Iwasaki, the Yale IMPACT Research Team, Richard A. Martinello, Albert I. Ko, Dana M. Small, Shelli F. Farhadian

## Abstract

**Background:** Healthcare workers (HCW) treating COVID-19 patients are at high risk for infection and may also spread infection through their contact with vulnerable patients. Smell loss has been associated with SARS-CoV-2 infection, but it is unknown whether monitoring for smell loss can be used to identify asymptomatic infection among high risk individuals, like HCW.

**Methods:** We performed a prospective cohort study, tracking 473 HCW across three months to determine if smell loss could predict SARS-CoV-2 infection in this high-risk group. HCW subjects completed a longitudinal, novel behavioral at-home assessment of smell function with household items, as well as detailed symptom surveys that included a parosmia screening questionnaire, and RT-qPCR testing to identify SARSCoV-2 infection.

**Results:** SARS-CoV-2 was identified in 17 (3.6%) of 473 HCW. Among the 17 infected HCW, 53% reported smell loss, and were more likely to report smell loss than COVID-negative HCW on both the at-home assessment and the screening questionnaire (*P* < .01). 67% reported smell loss prior to having a positive SARS-CoV-2 test, and smell loss was reported a median of two days before testing positive. Neurological symptoms were reported more frequently among COVID-positive HCW who reported smell loss (*P* < .01).

**Conclusions:** In this prospective study of HCW, self-reported changes in smell using two different measures were predictive of COVID-19 infection. Smell loss frequently preceded a positive test and was associated with neurological symptoms.

## INTRODUCTION

A salient feature of SARS-CoV-2, the virus that causes COVID-19, is its ability to rapidly spread, leading it to affect over 100 countries in a matter of weeks. It is increasingly clear that asymptomatic and pre-symptomatic infection play an important role in the ongoing spread of COVID-19 [1]. Peak infectiousness likely occurs on or before symptom onset [2, 3], with a significant proportion of secondary infections arising from the index case during the pre-symptomatic stage [4, 5], highlighting the urgent need for non-invasive screening tools to identify people who may be asymptomatic or pre-symptomatic. This is especially true for healthcare workers (HCW) treating COVID-19 patients. HCW are at high risk for nosocomial infection themselves and may unknowingly spread infection through contact with hospital staff and patients [6–8].

Reduced olfactory sensitivity, or anosmia, has been identified as a common manifestation of COVID-19. Olfactory loss has been identified by self-report in 15-85% of COVID-19 patients in Korea [9], Italy [8, 10–12], Spain [13], Canada [14, 15], the UK [16], and Iran [17]. Remarkably, the correlation between the risk of COVID-19 and self-reported olfactory loss has been reported to be as high as r = 0.87 [17]. Among patients with COVID-19 assessed for smell loss, 41% and 98% had impaired tests of olfaction on two different tests of olfactory function [18, 19]. Smell loss is also being evaluated for its diagnostic value in COVID-19 case identification [20]. One epidemiological study reported that average odor intensity ratings of household items predicted prevalence of COVID-19 in a Swedish population, with large differences in smell intensity ratings in individuals with and without COVID-19 symptoms [21]. Furthermore, individuals progressing from asymptomatic to reporting some COVID-19 symptoms had a large drop in odor intensity ratings. Another study reported loss of taste, smell, and chemesthesis following a COVID-19 diagnosis using a self-report questionnaire [22]. This compelling evidence highlights the potential importance of tracking smell sensitivity to identify SARS-CoV-2 infection. However, smell loss has not yet been prospectively evaluated in asymptomatic or pre-symptomatic individuals undergoing regular, frequent SARS-CoV-2 testing. Given the extreme infectiousness of the SARS-CoV-2 virus, it is critical to determine the onset and trajectory of smell loss, with respect to the time of infection, and whether smell loss predicts symptom type and/or severity.

Testing for objective smell loss using standard laboratory or clinical techniques is not feasible for widespread testing, and as self-report surveys are not sufficiently reliable, an accessible at-home smell sensitivity screen is of paramount importance. Smell-tracker at-home self-monitoring for the first sign of diminished smell function would enable rapid testing and/or self-quarantine, thus protecting the community from exposure and preventing further spread prior to a formal diagnosis.

The aim of this study was to determine if tracking smell sensitivity and loss using an at-home assessment could identify HCW who are infected with SARS-CoV-2. High risk HCW undergoing routine (every 3 days) viral screening for SARS-CoV-2 infection completed a novel at-home brief smell sensitivity screen, the Yale Jiffy (**Supplemental Table 1**), to track ratings of odor intensity perception using two household olfactory stimuli. HCW also self-reported smell loss and changes across this time, along with symptoms commonly used to screen for COVID-19.

**Table 1.** Baseline Characteristics of Participants in the Smell Sub-Study.

|  | COVID-19<br>Positive HCW<br>(*n* = 17) | COVID-19<br>Negative HCW<br>(*n* = 456) | *P*<br>Value<sup>a</sup> | Adjusted OR<br>(95% CI)<sup>b</sup> |
| --- | --- | --- | --- | --- |
| Demographics |  |  |  |  |
| Age, y | 30.0 (26.0, 48.0) | 34.5 (29.0, 44.0) | 0.47 | -- |
| Female sex | 15 (88) | 358 (79) | 0.54 | -- |
| Ethnicity |  |  | 0.84 | -- |
| White | 16 (94) | 359 (79) |  |  |
| Black | 0 (0) | 15 (3) |  |  |
| Hispanic | 0 (0) | 37 (8) |  |  |
| Asian | 1 (6) | 36 (8) |  |  |
| Other | 0 (0) | 9 (2) |  |  |
| BMI, kg/m<sup>2</sup> | 25.0 (22.2, 35.2) | 24.7 (22.7, 29.1) | 0.74 | -- |
| Occupation |  |  | 0.01 | -- |
| RN | 15 (88) | 246 (54) |  |  |
| MD | 0 (0) | 98 (21) |  |  |
| Other | 2 (12) | 112 (25) |  |  |
| Number of surveys<br>completed |  |  |  |  |
| Symptom survey | 10.0 (6.0, 22.0) | 22.0 (10.0, 34.0) | 0.08 | -- |
| Yale Jiffy<sup>c</sup> | 1.5 (1.0, 4.5) | 8 (2.0, 24.0) | 0.03 | -- |
| Smell loss |  |  |  |  |
| Yale Jiffy | 5/9 (56) | 43/304 (14) | 0.005 | -- |
| Symptom survey | 8/17 (47) | 83/456 (18) | 0.008 | -- |
| Either survey | 9/17 (53) | 105/456 (23) | 0.009 | 4.52 (1.61, 13.3) |

Data are presented as median (IQR) for continuous variables and no. (%) for categorical variables.
Abbreviations: BMI, body mass index; CI: confidence interval; HCW, healthcare workers; MD, medical doctor; OR, odds ratio; RN, registered nurse.
<sup>a</sup> Unadjusted *P* values are Wilcoxon rank sum test (continuous variables) or Fisher's exact test (categorical variables).
<sup>b</sup> Adjusted odds ratios and 95% confidence intervals for symptoms were obtained from separate logistic regression models, each adjusted for age, sex, BMI, and number of symptom surveys completed.
<sup>c</sup> *n* = 9 for COVID-19 positive HCW; *n* = 304 for COVID-19 negative HCW.

## METHODS

### Study Setting, Population, and Recruitment

We conducted a prospective cohort study of smell symptomology nested within the Implementing Medical and Public Health Action against Coronavirus (CT) (IMPACT) study at Yale University (HIC # 200027690). The goal of the parent study was to prospectively follow COVID-negative HCW at high risk of acquiring infection due to occupational exposures. The study recruited HCW working in the medical ICU or dedicated COVID-19 units at Yale New Haven Hospital (YNHH), a 1,541 bed tertiary care hospital located in New Haven, CT, USA. All participants provided written and/or verbal informed consent. Inclusion criteria for the IMPACT study included: a) aged 18 or older; b) English-speaking; c) working in a health care facility (YNHH, Yale Health); d) possible moderate to high risk exposure to COVID-19, or work in a COVID-19 unit, and e) SARS-CoV-2 negative at study entry. For this analysis of smell alterations and COVID-19, we excluded subjects without at least one SARS-CoV-2 PCR result and those who had not completed at least one daily symptom questionnaire or Yale Jiffy. All reported data were collected between March 31 and July 7, 2020.

### Viral testing

SARS-CoV-2 real-time quantitative polymerase chain reaction (RT-qPCR) testing was performed on self-collected nasopharyngeal and saliva specimens every three days. Specimens were processed and tested on the same day as collection following previously described protocols [23, 24]. Two HCW reported positive results from a CLIA-certified lab outside of the study protocol.

### Yale Jiffy

The Yale Jiffy is an online survey developed to screen for smell loss that can be conducted in under two minutes using readily available household items. The questionnaire includes two sections using self-ratings of: 1) ability to smell, and 2) strength of smell in response to olfactory and trigeminal stimuli. Peanut butter (or jam/jelly) was used as the olfactory stimulus as it has minimal or no trigeminal component, allowing for isolation of effects on the olfactory system. We also included a stimulus with a trigeminal component as a control stimulus (i.e., vinegar).

First, participants were asked to rate their ability to smell on a categorical scale (Poor/Average/Good/Very Good), to report any reduction in smell in the past week on a categorical scale (None/Slight/Moderate/Severe), and to rate the degree of reduction on a 10cm (0.0-10.0) visual analog scale (VAS). Next, participants were asked to hold the olfactory stimulus one inch from their nose and to provide ratings of both the strength of smell (0.0-10.0) and how different it smells from normal (0.0-10.0). HCW then held the trigeminal stimulus one inch from their nose and rated strength of sensation of irritation and difference from normal, as above. Since olfactory sensitivity fluctuates across the day [25], HCW were asked to complete the survey at approximately the same time each day using the same stimuli each time they completed the test.

Responses were collected using Qualtrics, a secure HIPAA-compliant web-based survey platform, and retention was encouraged using daily e-mail reminders.

### Daily Symptom Questionnaire

As part of the IMPACT study, HCW completed an online daily symptom questionnaire. HCW were given a list of symptoms and prompted to indicate whether they had developed such symptoms in the past 24 hours. Listed symptoms included objective fever (≥100.4°F), subjective fever, cough, shortness of breath, stuffy nose, sore throat, chills, sweating, malaise, fatigue, muscle pain, anorexia, nausea, vomiting, diarrhea, abdominal pain, and dizziness. In addition, we included screening questions for parosmia – changes in odor quality perception [26] – and for hypogeusia – reduced ability to taste [27]. Participants completed four parosmia screening questions (**Supplemental Table 2**), indicating how often they were bothered by common complaints caused by smell distortions, with responses ranging from “always” (1 point) to “never” (4 points). Parosmia was defined as a cumulative score less than or equal to 14 (out of 16). HCW responded to four hypogeusia screening questions for saltiness, sourness, sweetness, and bitterness, by indicating that they could detect these tastes “Easily” (3 points), “Somewhat” (2 points), or “Not at all” (1 point). Hypogeusia was defined as any total score less than the maximum of 12 points.

**Table 2.** Characteristics of COVID-19 Positive Healthcare Workers by Reported Smell Change.

|  | Smell Change<br>(*n* = 9) | No Smell Change<br>(*n* = 8) | *P* Value<sup>a</sup> |
| --- | --- | --- | --- |
| Cycle threshold, mean (SD) | 27.7 (4.9) | 30.2 (3.5) | 0.28 |
| Age | 34 (30, 59) | 26.5 (25.8, 33.8) | 0.054 |
| Female sex | 9 (100) | 6 (75.0) | 0.21 |
| Non-white race | 0 (0) | 1 (12.5) | 0.47 |
| BMI | 26.6 (22.3, 35.5) | 23.7 (22.6, 27.7) | 0.71 |
| Smell change as presenting symptom | 3 (33.3) | -- | -- |
| Any neurological symptoms<sup>b</sup> | 9 (100) | 3 (37.5) | 0.009 |
| Neurological symptoms ≥ 7 days after positive test<sup>b</sup> | 4 (44.4) | 1 (12.5) | 0.29 |

Data are presented as median (IQR) for continuous variables and no. (%) for categorical variables unless otherwise indicated.
Abbreviations: BMI, body mass index.
<sup>a</sup> Unadjusted *P* values obtained from student's *t* test (normally distributed continuous variables), Wilcoxon rank sum test (non-normally distributed continuous variables), and Fisher's exact test (categorical variables).
<sup>b</sup> Neurological symptoms include headache, dizziness, and fatigue.

### Statistical Analyses

Descriptive statistics were used to characterize the study population. The Fisher’s exact and Wilcoxon rank-sum tests were used to compare COVID-19-positive and negative HCW. Multivariable logistic regression models were developed to calculate adjusted odds ratios for the associations between smell symptoms and COVID-19 diagnosis. Each model included one symptom as the predictor and additionally adjusted for age, sex, body mass index (BMI), ethnicity, and number of symptom surveys completed.

To further examine the strength of association between symptoms of smell loss and a diagnosis of COVID-19, we compared HCW reporting smell loss to a subset of HCW never reporting smell loss who were well-matched on age, sex, ethnicity, and the number of daily symptom surveys completed. We used the R package ‘MatchIt’ for this analysis, specifying a control to case ratio of 2:1 and employing optimal matching, which minimizes the overall differences between cases and controls. To estimate the strength of association between smell loss and a diagnosis of COVID-19, we conducted conditional logistic regression using the matched dataset and the R package ‘survival.’ We compared results of three matched logistic regression analyses among those reporting smell loss on either survey (Yale Jiffy or daily symptom questionnaire), on the Yale Jiffy only, and on the daily symptom questionnaire only. Sensitivity analyses tested whether inclusion of covariates was needed to adjust for residual confounding after matching, determined by a change in the point estimate of > 10%.

Finally, we summarized responses to the Yale Jiffy and daily symptom questionnaires and used Fisher’s exact and Wilcoxon rank-sum tests to compare responses between COVID-19-positive and COVID-19-negative HCW. We also examined longitudinal responses among HCW who completed the questionnaires multiple times.

All analyses were conducted with a two-sided statistical significance level of *P* < .05 using R statistical software (version 3.4.2).

## RESULTS

### Participant characteristics

588 HCW were recruited and consented in the IMPACT study between March 31 and July 7, 2020, of whom 473 (80%) were eligible for the smell sub-study (**Figure 1**). Within the sub-study population, 373 (79%) participants were female; the mean (SD) age was 37.5 (11.2) years; 375 (79%) were white non-Hispanic/Latino; 261 (55%) were registered nurses (RNs) and 98 (21%) were medical doctors (MDs) (**Table 1**). Compared to HCW included in this analysis, IMPACT HCW ineligible for the sub-study were more likely to be Black, Asian, or other ethnicity (*P* < .001) and have higher BMI (*P* = 0.004) (**Supplemental table 3**).

**Figure 1:**
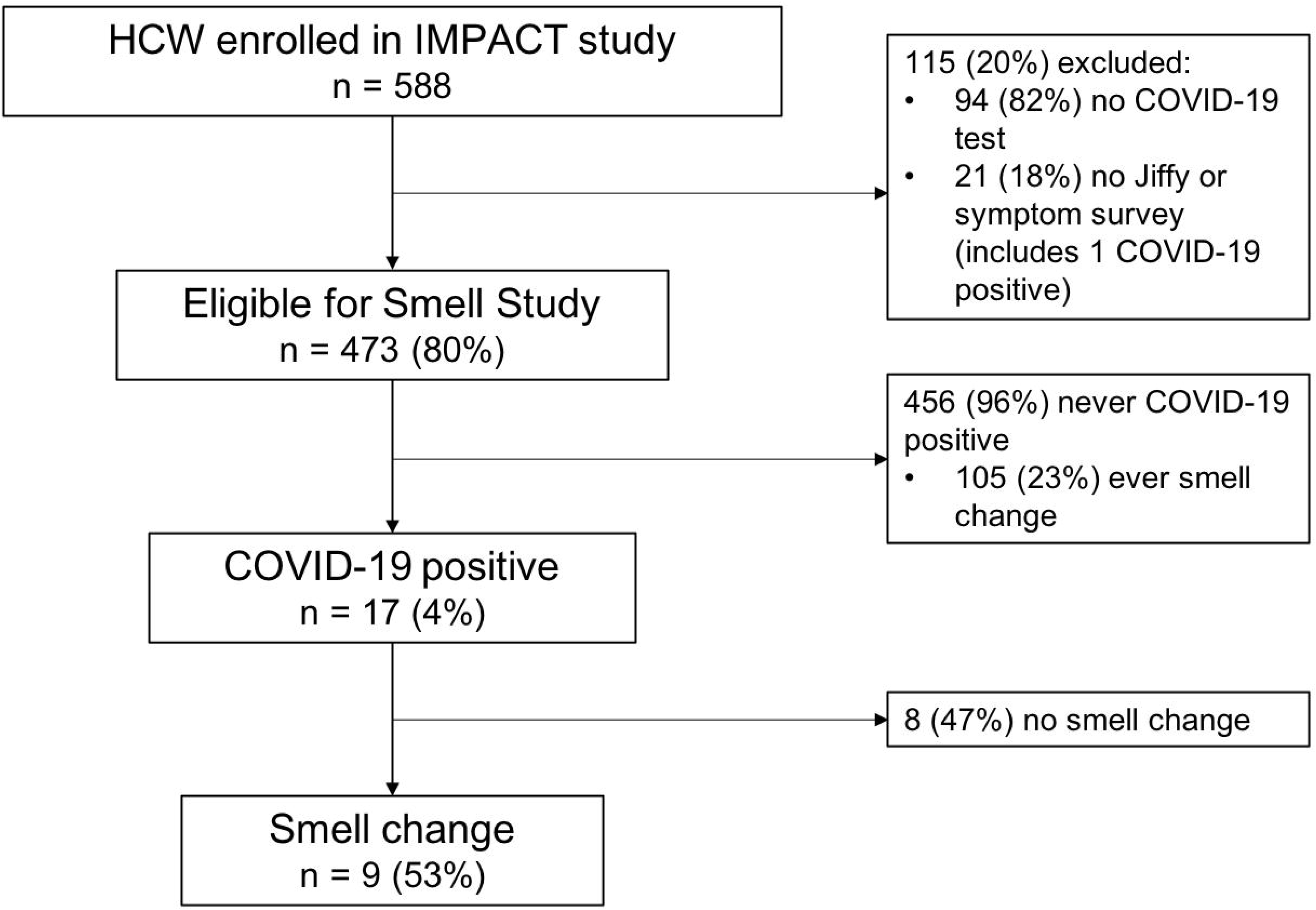
Flow diagram of participants and reported changes in smell by COVID-19 status. Abbreviations: HCW, healthcare workers; IMPACT, Implementing Medical and Public Health Action against Coronavirus (CT).

HCW included in the sub-study completed the daily symptom questionnaire (including parosmia/hypogeusia screening) a median 23 times per HCW (IQR: 10, 34). Among the 313 (66%) HCW who completed the Yale Jiffy at least once, HCW completed a median of 10 (IQR: 3, 28) Jiffy questionnaires.

5771 SARS-CoV-2 RT-qPCR tests were performed on 473 HCW in the sub-study (median = 11 tests per HCW) between March 31 and July 7. Of these, 17 (3.6%) HCW tested positive for SARS-CoV-2.

### HCW who eventually tested positive for COVID-19 were more likely to report smell loss on the Yale Jiffy and daily symptom questionnaire

The demographic characteristics of COVID-positive HCW were similar to those of COVID-19-negative HCW (**Table 1**). Nine of the 17 (53%) COVID-positive HCW completed the Yale Jiffy at least once, but most began rating the household items only after they reported categorical smell loss. Therefore, the number of individuals with a positive diagnosis and pre-post ratings was too small to analyze for smell loss. COVID- positive HCW were more likely to report categorical smell loss on the Jiffy, with 5/9 (56%) COVID-positive HCW versus 43/304 (14%) COVID-negative HCW reporting smell loss (OR = 7.6, 95% CI: 2.0-29.4) (**Table 1**). The five COVID-positive HCW who reported categorical smell loss via the Jiffy had a mean (SD) reduction in smell of 5.8 (4.0)cm on a 10cm (0.0 to 10.0) scale; those indicating severe loss had a mean 8.3 (3.0)cm decrease compared to 2.2 (0.5)cm in those with slight smell loss. For individuals with any smell loss reported via Jiffy, COVID-positive HCW reported more severe smell loss than COVID-negative HCW (*P* < .001) (**Figure 2**); COVID-positive HCW reported severe (60%) or slight (40%) smell loss on a categorical scale, while COVID-negative HCW reported slight (88%) or moderate (12%) smell loss. Pairwise comparisons at each level of smell loss severity showed that COVID-positive HCW were more likely to report severe smell loss compared to COVID-negative HCW reporting any other levels of smell loss (*Ps* < .05).

**Figure 2:**
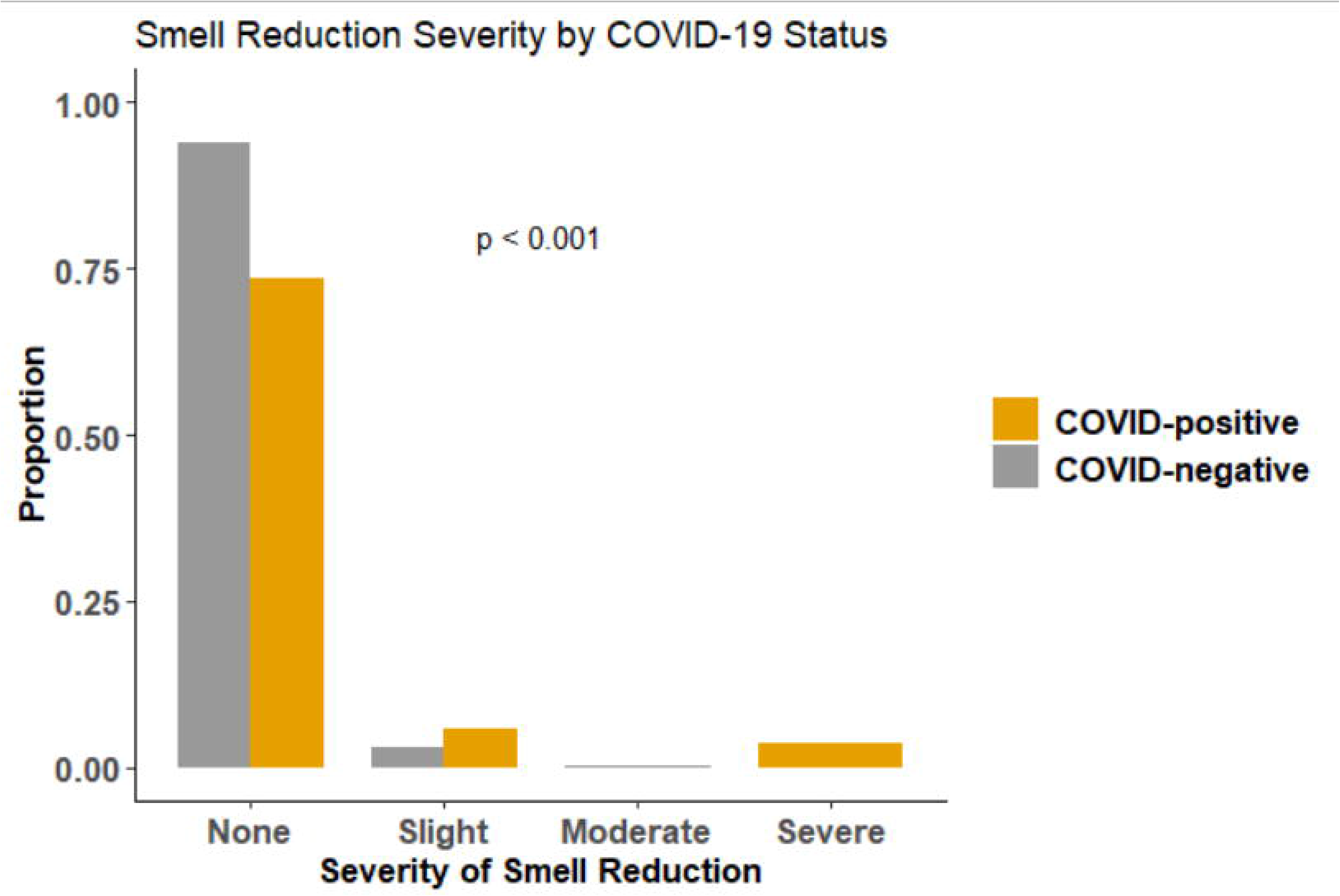
Comparison of the proportions of self-reported severity of smell loss on the Yale Jiffy by COVID-positive and COVID-negative healthcare workers. *P* value is a Fisher’s exact test of independence.

All 17 COVID-positive HCW eligible for the smell sub-study completed at least one daily symptom questionnaire. Eight (47%) reported parosmia versus 83/456 (18%) COVID-negative HCW (OR = 4.0, 95% CI: 1.5-10.7). Hypogeusia was reported in seven (41%) COVID-positive HCW and 104/456 (23%) COVID-negative HCW (OR = 2.4, 95% CI: 0.7, 7.1) (**Table 1**). Six of those seven also reported smell loss.

Relative to Day 0 (defined as the day of test positivity), the median timing of reported smell loss was Day −2 (IQR, Day −16 to Day +2) among COVID-positive HCW reporting smell loss, with 6/9 (67%) reporting smell loss before test positivity (**Figure 3**). Subjects recovered their sense of smell a median of nine days (IQR, 4, 16.5) after first reporting smell loss. Of those who reported smell loss, 3/9 (33%) reported parosmia or smell loss as their first symptom.

**Figure 3:**
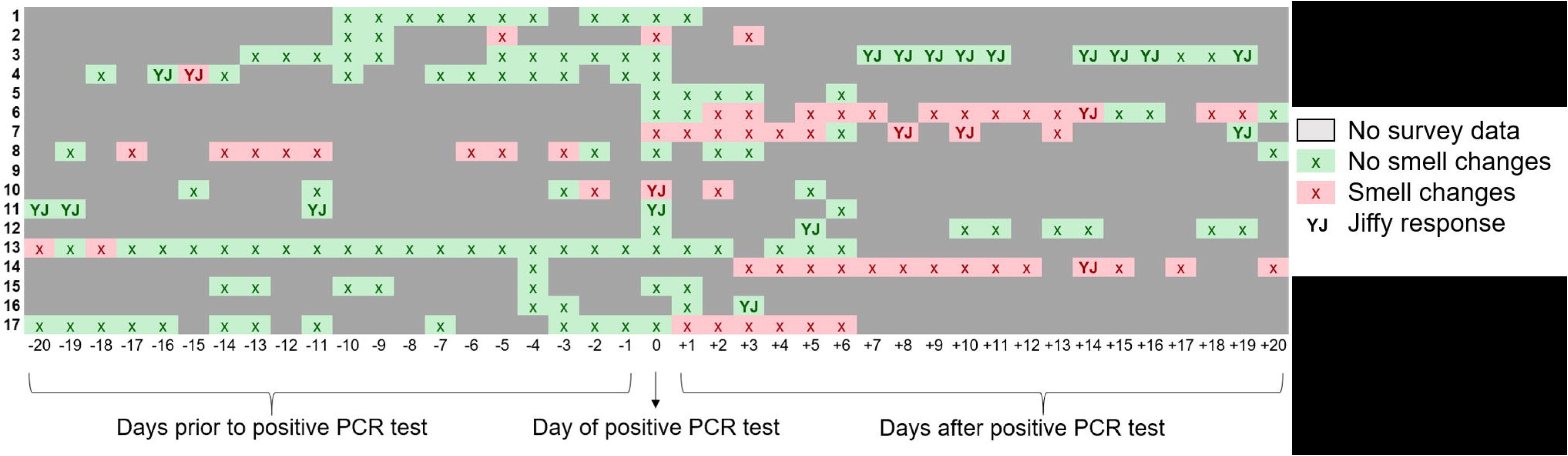
Chronology of smell changes among COVID-positive healthcare workers relative to day of positive test. Red boxes indicate smell change was reported via either the symptom survey or Yale Jiffy. Green boxes indicate no smell change reported. Solid gray boxes indicate there was no Yale Jiffy or symptom survey submitted for that day. Boxes with a “YJ” specifically indicate a Yale Jiffy response.

### Associations between smell symptoms and COVID-19 diagnosis

Overall, in either the daily symptom questionnaire or Yale Jiffy, nine (53%) COVID-positive HCW reported smell loss or parosmia, compared to 105/456 (23%) COVID-negative HCW (OR = 3.7, 95% CI: 1.2, 11.5). After adjusting for age, sex, BMI, and number of symptom questionnaires, smell loss remained a significant predictor of a COVID-19 diagnosis (aOR = 4.5, 95% CI: 1.6, 13.3). Among other symptoms in the questionnaire, only dyspnea (aOR 7.3, 95% CI: 2.2, 22.4) and headache (aOR 6.1, 95% CI: 1.8, 28.4) were more strongly predictive of SARS-CoV-2 infection.

Likewise, in the matched analysis, 114 HCW reported any smell loss in either the daily symptom questionnaire or the Yale Jiffy. Optimal matching resulted in a sample of 228 HCW who never reported smell loss. The results of matching on age, sex, ethnicity, BMI, and number of questionnaires are shown in **Supplemental Figure 1**. Using this matched dataset, conditional logistic regression estimated a significant association between smell loss and SARS-CoV-2 infection (OR = 6.0, 95% CI 1.6-22.2). Similar results were obtained when restricting the matched analysis to 89 HCW reporting smell loss via the daily symptom questionnaire only (178 matched controls, OR = 3.2, 95% CI 1.1-9.8) or the 47 HCW who reported smell loss via the Yale Jiffy only (94 matched controls, OR = 5.0, 95% CI: 0.97-25.8), noting the smaller sample size for the Yale Jiffy resulted in a less precise estimate. Results were not sensitive to the inclusion of covariates to adjust for residual confounding after matching.

### Associations between smell loss and neurological symptoms in COVID-19 positive HCW

**Table 2** summarizes findings among COVID-positive HCW stratified by smell loss as reported by either measure. COVID-positive HCW who reported smell loss were older than those who did not report any smell loss (median [IQR], 35.5 [29.8, 59.8] vs. 26.0 [25.0, 29.0] years, *P* = 0.05). There was also a trend toward lower cycle threshold among COVID-positive HCW reporting parosmia specifically (mean [SD], 26.8 [4.5] vs. 30.9 [3.2], *P* = 0.07), but not among HCW reporting any smell changes in either survey. Neurological symptoms as assessed by the daily symptom questionnaire, including headache, fatigue, and dizziness, were reported in all nine COVID-positive HCW reporting smell loss, and three (38%) who did not report smell loss. Prolonged neurological symptoms (> seven days after positive test) were reported in four (44%) COVID-positive HCW with smell loss versus one (13%) without smell loss. Three COVID-positive HCW, all of whom experienced smell loss, had significantly prolonged neurological symptoms (> 20 days after positive test).

## DISCUSSION

In this prospective study of a high-risk HCW population, we assessed smell loss alongside routine viral testing, finding that HCW who acquired SARS-CoV-2 infection over the course of the study had significantly increased odds of reporting smell loss compared to COVID-negative HCW. This finding was consistent across bivariate (OR = 3.7), regression-adjusted (OR = 4.5), and matched (OR = 6.0) analyses. Likewise, COVID-positive HCW reported significantly more severe smell loss compared to COVID-negative HCW. Overall, our findings add to the accumulating evidence demonstrating the efficacy and feasibility of using prospective, self-administered, at-home assessments of smell sensitivity to track changes over time in a group of individuals at high risk for SARS-CoV-2 infection.

Our findings are consistent with recent research demonstrating robust associations between smell perturbations and SARS-CoV-2 infection. Specifically, smell and taste impairments are reported prior to (20%) or during (13%) hospitalization [10], typically lasting 4-17 days [28], and provide better predictive ability than fever or cough [29], two of the more commonly used symptoms to screen for COVID-19. Indeed, a large cross-sectional study found that sudden self-reported smell loss was the best predictive indicator of COVID-19 infection [30]. Our study extends these findings by uniquely assessing for prospective smell loss alongside viral screening in a high-risk group, all of whom were COVID-19 negative at study entry. Moreover, distortion in smell was a better predictor of COVID-19 than many other physical symptoms. Cough, headache, and dyspnea were also significantly associated with increased odds of COVID-19 diagnosis in this study. However, these symptoms are commonly experienced in other respiratory infections and not specific symptoms for COVID-19, and therefore less useful than loss of smell for monitoring for COVID-19 among asymptomatic individuals.

Interestingly, we found an association between reduction in smell ability and neurological symptoms in COVID-19. While the exact pathophysiology underlying COVID-19 related smell loss remains incompletely understood, the association between smell loss and other neurological symptoms suggests a potential common mechanism underlying these symptoms. A growing body of evidence has demonstrated the potential for SARS-CoV-2 to infect neurons [31, 32], and our results lend support to the hypothesis that smell loss may be related to central nervous system infection by SARSCoV-2, consistent with studies reporting MRI abnormalities in the olfactory bulb in a subset of patients with COVID-19 related anosmia [33, 34].

Our study has several limitations. Smell sensitivity and loss using the Yale Jiffy were evaluated using both categorical responses and a VAS [35]. Indeed, those indicating severe loss reported an average of an 8.3cm decrease on the 10cm scale, compared to a 2.2cm reduction in those indicating slight loss, suggesting categorical responses are sufficient for identifying asymptomatic carriers. However, because many participants already had smell loss the first time they completed the VAS, we were not able to detect any changes. Consistent daily ratings of a household item could prove useful for further evaluating the magnitude of loss but would require further instructions and possibly practice using the scale. Adopting the generalized labeled magnitude scale (gLMS), a specialized line scale with semantic labels at empirically derived intervals, may also improve outcomes, as this scale is less subject to floor and ceiling effects [36]. However, it requires some training and practice. Additionally, the low number of positive cases limited statistical power.

In our study, self-reported changes in smell perception were predictive of SARSCoV-2 infection in a healthcare worker population. At-home smell assessments should be considered for non-invasive screening of groups that are at high risk for COVID-19.

## Data Availability

All data generated and/or analysed during the current study are available from the corresponding author on reasonable request.

## Notes

### Yale IMPACT Research Team authors

(Listed in alphabetical order) Staci Cahill, Edward Courchaine, Christina Harden, Chaney Kalinch, Daniel Kim, Lynda Knaggs, Eriko Kudo, Peiwen Lu, Alice Lu-Culligan, Nida Naushad, Allison Nelson, Isabel M. Ott, Annsea Park, Mary Petrone, Sarah Prophet, Lorenzo Sewanan, Maria Tokuyama, Jordan Valdez, Arvind Venkataraman, Chantal B.F. Vogels, Annie Watkins, Yexin Yang

## Acknowledgements

The authors are grateful to the study participants for their time and commitment to the study. We thank all members of the staff at the Yale IMPACT Study and to Mr. Brett Wilson for his generous donation to support this project.

## References

1. Bai Y, Yao L, Wei T, et al. Presumed Asymptomatic Carrier Transmission of COVID-19. JAMA 2020; 323(14): 1406–7.

2. He X, Lau EHY, Wu P, et al. Temporal dynamics in viral shedding and transmissibility of COVID-19. Nature Medicine 2020; 26(5): 672–5.

3. Furukawa N, Brooks J, Sobel J. Evidence Supporting Transmission of Severe Acute Respiratory Syndrome Coronavirus 2 While Presymptomatic or Asymptomatic. Emerging Infectious Disease journal 2020; 26(7).

4. Furuse Y, Sando E, Tsuchiya N, et al. Clusters of Coronavirus Disease in Communities, Japan, January–April 2020. Emerging Infectious Disease journal 2020; 26(9): 2176.

5. Du Z, Xu X, Wu Y, Wang L, Cowling B, Meyers LA. Serial Interval of COVID-19 among Publicly Reported Confirmed Cases. Emerging Infectious Disease journal 2020; 26(6): 1341.

6. Nguyen LH, Drew DA, Graham MS, et al. Risk of COVID-19 among front-line health-care workers and the general community: a prospective cohort study. The Lancet Public Health 2020; 5(9): e475–e83.

7. Lai X, Wang M, Qin C, et al. Coronavirus Disease 2019 (COVID-2019) Infection Among Health Care Workers and Implications for Prevention Measures in a Tertiary Hospital in Wuhan, China. JAMA Network Open 2020; 3(5): e209666–e.

8. Piapan L, De Michieli P, Ronchese F, et al. COVID-19 outbreak in healthcare workers in Trieste hospitals (North-Eastern Italy). J Hosp Infect 2020: S0195-6701(20)30390-X.

9. Lee Y, Min P, Lee S, Kim S-W. Prevalence and Duration of Acute Loss of Smell or Taste in COVID-19 Patients. J Korean Med Sci 2020; 35(18).

10. Giacomelli A, Pezzati L, Conti F, et al. Self-reported Olfactory and Taste Disorders in Patients With Severe Acute Respiratory Coronavirus 2 Infection: A Cross-sectional Study. Clinical Infectious Diseases 2020; 71(15): 889–90.

11. Spinato G, Fabbris C, Polesel J, et al. Alterations in Smell or Taste in Mildly Symptomatic Outpatients With SARS-CoV-2 Infection. JAMA 2020; 323(20): 2089–90.

12. Vaira LA, Deiana G, Fois AG, et al. Objective evaluation of anosmia and ageusia in COVID-19 patients: Single-center experience on 72 cases. Head & Neck 2020; 42(6): 1252–8.

13. Gómez-Iglesias P, Porta-Etessam J, Montalvo T, et al. An Online Observational Study of Patients With Olfactory and Gustory Alterations Secondary to SARSCoV-2 Infection. Front Public Health 2020; 8: 243.

14. Carignan A, Valiquette L, Grenier C, et al. Anosmia and dysgeusia associated with SARS-CoV-2 infection: an age-matched case–control study. Canadian Medical Association Journal 2020; 192(26): E702–E7.

15. Lee DJ, Lockwood J, Das P, Wang R, Grinspun E, Lee JM. Self-reported anosmia and dysgeusia as key symptoms of coronavirus disease 2019. CJEM 2020: 1–8.

16. Patel A, Charani E, Ariyanayagam D, et al. New-onset anosmia and ageusia in adult patients diagnosed with SARS-CoV-2 infection. Clin Microbiol Infect 2020; 26(9): 1236–41.

17. Bagheri SH, Asghari A, Farhadi M, et al. Coincidence of COVID-19 epidemic and olfactory dysfunction outbreak in Iran. Medical Journal of the Islamic Republic Of Iran 2020; 34(1): 446–52.

18. Hornuss D, Lange B, Schröter N, Rieg S, Kern WV, Wagner D. Anosmia in COVID-19 patients. Clin Microbiol Infect 2020: S1198-743X(20)30294-9.

19. Moein ST, Hashemian SM, Mansourafshar B, Khorram-Tousi A, Tabarsi P, Doty RL. Smell dysfunction: a biomarker for COVID-19. International Forum of Allergy & Rhinology 2020; 10(8): 944–50.

20. Haehner A, Draf J, Dräger S, de With K, Hummel T. Predictive Value of Sudden Olfactory Loss in the Diagnosis of COVID-19. ORL J Otorhinolaryngol Relat Spec 2020; 82(4): 175–80.

21. Iravani B, Arshamian A, Ravia A, et al. Relationship Between Odor Intensity Estimates and COVID-19 Prevalence Prediction in a Swedish Population. Chem Senses 2020; 45(6): 449–56.

22. Parma V, Ohla K, Veldhuizen MG, et al. More than smell – COVID-19 is associated with severe impairment of smell, taste, and chemesthesis. Chem Senses 2020

23. Wyllie AL, Fournier J, Casanovas-Massana A, et al. Saliva or Nasopharyngeal Swab Specimens for Detection of SARS-CoV-2. New England Journal of Medicine 2020

24. Takahashi T, Ellingson MK, Wong P, et al. Sex differences in immune responses that underlie COVID-19 disease outcomes. Nature 2020.

25. Herz RS, Van Reen E, Barker DH, Hilditch CJ, Bartz AL, Carskadon MA. The Influence of Circadian Timing on Olfactory Sensitivity. Chem Senses 2017; 43(1): 45–51.

26. Landis BN, Frasnelli J, Croy I, Hummel T. Evaluating the clinical usefulness of structured questions in parosmia assessment. Laryngoscope 2010; 120(8): 1707–13.

27. Soter A, Kim J, Jackman A, Tourbier I, Kaul A, Doty RL. Accuracy of Self-Report in Detecting Taste Dysfunction. The Laryngoscope 2008; 118(4): 611–7.

28. Levinson R, Elbaz M, Ben-Ami R, et al. Time course of anosmia and dysgeusia in patients with mild SARS-CoV-2 infection. Infectious Diseases 2020; 52(8): 600℃2.

29. Menni C, Valdes AM, Freidin MB, et al. Real-time tracking of self-reported symptoms to predict potential COVID-19. Nature Medicine 2020; 26(7): 1037–40.

30. Gerkin RC, Ohla K, Veldhuizen MG, et al. The best COVID-19 predictor is recent smell loss: a cross-sectional study. medRxiv 2020: 2020. 07.22.20157263.

31. Zhang B-Z, Chu H, Han S, et al. SARS-CoV-2 infects human neural progenitor cells and brain organoids. Cell Research 2020

32. Solomon IH, Normandin E, Bhattacharyya S, et al. Neuropathological Features of Covid-19. New England Journal of Medicine 2020; 383(10): 989–92.

33. Laurendon T, Radulesco T, Mugnier J, et al. Bilateral transient olfactory bulb edema during COVID-19–related anosmia. Neurology 2020; 95(5): 224–5.

34. Politi LS, Salsano E, Grimaldi M. Magnetic Resonance Imaging Alteration of the Brain in a Patient With Coronavirus Disease 2019 (COVID-19) and Anosmia. JAMA Neurology 2020; 77(8): 1028–9.

35. Bartoshuk LM, Duffy VB, Fast K, Green BG, Prutkin J, Snyder DJ. Labeled scales (e.g., category, Likert, VAS) and invalid across-group comparisons: what we have learned from genetic variation in taste. Food Quality and Preference 2003; 14(2): 125–38.

36. Bartoshuk LM, Duffy VB, Green BG, et al. Valid across-group comparisons with labeled scales: the gLMS versus magnitude matching. Physiology & Behavior 2004; 82(1): 109–14.

